# Cost-effectiveness analysis of COVID-19 mRNA XBB.1.5 Fall 2023 vaccination in the Netherlands

**DOI:** 10.1101/2024.09.26.24314420

**Authors:** Florian Zeevat, Simon van der Pol, Tjalke Westra, Ekkehard Beck, Maarten J. Postma, Cornelis Boersma

## Abstract

**Objective:** This study aims to assess the cost-effectiveness of the Fall 2023 COVID-19 mRNA XBB.1.5 vaccination campaign in the Netherlands, comparing the XBB1.5 updated mRNA-1273.222 with the XBB1.5 updated BNT162b2 vaccine.

**Methods:** A one-year decision tree-based cost-effectiveness model was developed, considering three scenarios: no Fall 2023 vaccination, BNT162b2 vaccination and mRNA-1273 vaccination in the COVID-19 high-risk population in the Netherlands. The high-risk population includes everyone of 60 and older, and younger adults at high risk as identified by the Dutch Health Council. Costs were included from a societal perspective and the modelled period started in October 2023 and ended in September 2024, including life years lost with a lifetime horizon. Sensitivity and scenario analyses were conducted to evaluate model robustness.

**Results:** In the base case, mRNA-1273 demonstrated substantial benefits over BNT162b2, potentially averting 20,629 symptomatic cases, 924 hospitalizations (including 32 ICU admissions), 207 deaths, and 2,124 post-COVID cases. Societal cost savings were €12.9million (excluding vaccination costs), with 1,506 QALYs gained. The break-even incremental price of mRNA-1273 compared to BNT162b2 was €16.72 or €34.32 considering a willingness to pay threshold (WTP) of 20,000 or 50,000 euro per QALY gained.

**Conclusion:** This study provides a comprehensive cost-effectiveness analysis supporting the adoption of the mRNA-1273 vaccine in the national immunization programme in the Netherlands, provided that the Dutch government negotiates a vaccine price that is at most €34.32 per dose higher than BNT162b2. Despite limitations, the findings emphasize the substantial health and economic benefits of mRNA-1273 over BNT162b2 in the high-risk population.

## Introduction

The COVID-19 pandemic has been a global health crisis of unprecedented proportions, profoundly impacting public health systems and economies across the world^1^. Since the onset of the pandemic, vaccines have been the most effective intervention in preventing the spread of the SARS-CoV-2 virus and reducing the clinical burden of COVID-19^2^. Currently, SARS-CoV-2, particularly Omicron and its subvariants, reached an endemic stage in the Netherlands^3^.

Despite the substantial levels of COVID-19 vaccination coverage and immunity resulting from prior infections and vaccinations, the emergence of the Omicron variant in late 2021 triggered a rapid surge in COVID-19 cases, affecting various countries, including the Netherlands. The implementation of a proactive strategy has been critical for maintaining and strengthening immunity across the population. This strategy included the introduction of booster vaccinations with the aim of increasing the durability and effectiveness of protection against COVID-19.

The Netherlands launched its COVID-19 booster vaccination campaign in November 2021, initially targeting those aged 18 years and older. By February 2022, eligibility for a second COVID-19 booster was limited to individuals at higher risk, such as those aged 70 and older, residents of nursing homes, adults with Down syndrome, and those with severe immunodeficiency, provided they had completed both their initial vaccination series and the booster shot^4^. On September 19, 2022, a new phase of the booster campaign was launched, featuring the use of bivalent COVID-19 mRNA vaccines, namely BNT162b2 (Comirnaty, BioNTech/Pfizer) and mRNA-1273.222 (Spikevax, Moderna)^5^. The Minister of the Netherlands decided, in September 2023, to initiate a structural vaccination program, on the advice of the Health Council^6,7^. The public vaccination programme were extended to various target groups, including individuals aged 60 and older, medical high-risk groups, pregnant women, and healthcare workers^8^. In October 2023, a new round of vaccinations started, using the BNT162b2 monovalent XBB.1.5 vaccine to target the Omicron XBB lineage that was at the time predominant, thus replacing the previous bivalent vaccine.

As of April 2023, mRNA-1273 is no longer part of the COVID-19 vaccination programme in the Netherlands; only the monovalent BNT162b2 is being administered^9^, despite both mRNA technology-based vaccines being licensed in Netherlands. The vaccines differ in delivery system and dosage, with the mRNA-1273 vaccine containing a higher dosage of active ingredient (50 µg for the booster) compared to the BNT162b2 vaccine (30 µg of mRNA for the booster). These differences may impact vaccine effectiveness, as demonstrated in high-risk patients such as older adults, where a meta-analysis considering studies evaluating both primary series and booster of mRNA-1273 (100 and 50 µg) showed lower risk compared to BNT162b2 (30 µg) in SARS-CoV-2 infections (risk ratio 0.72) and COVID-19-related hospital admissions (risk ratio 0.65)^10^.

The aim of this study was to estimate the cost-effectiveness of a Fall 2023 COVID-19 booster campaign with the XBB.1.5 updated mRNA-1273 vaccine compared to the XBB1.5 BNT162b2 vaccine in the Netherlands. This can inform the decisions regarding COVID-19 vaccination in 2024 and beyond.

## Methods

### Model overview

A decision tree-based cost-effectiveness model (Figure 1) was developed^11^ to estimate the cost-effectiveness of an XBB1.5 updated mRNA-1273.222 vaccination programme from a societal perspective. The analysis covered a one-year time horizon (e.g., October-September), during which the long-term effects of COVID were also accounted for. The primary comparison focused on the XBB1.5 BNT162b2, which is the current standard in the Netherlands, but no vaccination was also included in the analysis. The XBB1.5 updated vaccines will be referred to as mRNA-1273 and BNT162b2 in the remainder of this manuscript. The model simulates the consequences of infections, including resource utilization, mortality, and their impact on quality of life. The model incorporates the short-term “infection period” and covers immediate infection-related consequences, including infection-related myocarditis, outpatient care, hospitalizations including ICU, and post-hospitalization recovery as well as the “post-COVID” period accounting for any long-term impacts resulting from the COVID-19 infection.

**Figure 1.**
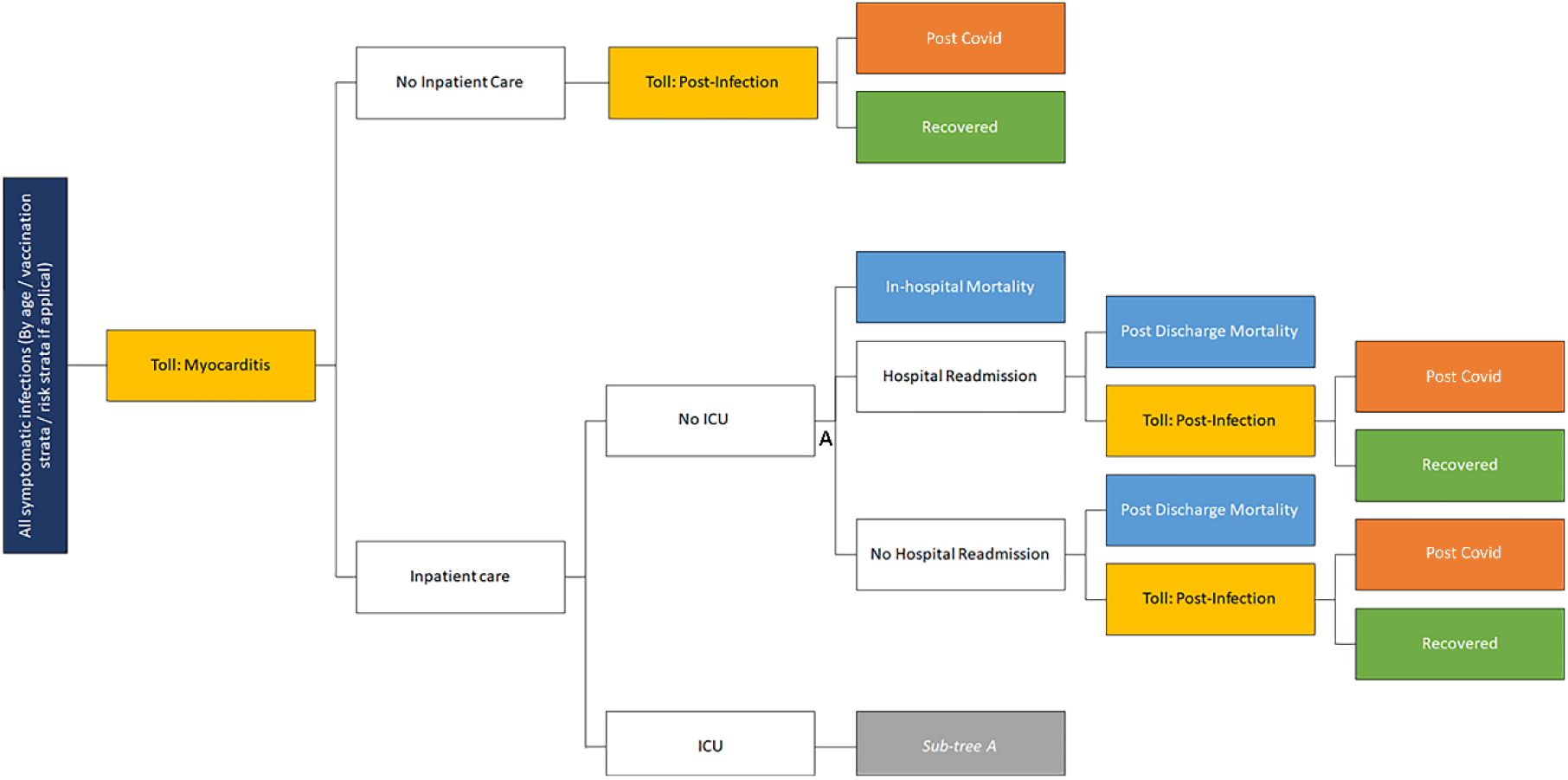
Overview of the full model structure. ICU: Intensive Care Unit; Toll: The outcome does not occur in all cases, but in a percentage of the patients.

### Model Input

All input parameters of the model can be found in Table 1. Costs were expressed in 2023 price levels and were not discounted given that the model’s time horizon is limited to one year. However, health effects associated with premature death were considered beyond one year and discounted with 1.5%^12^.

**Table 1.**
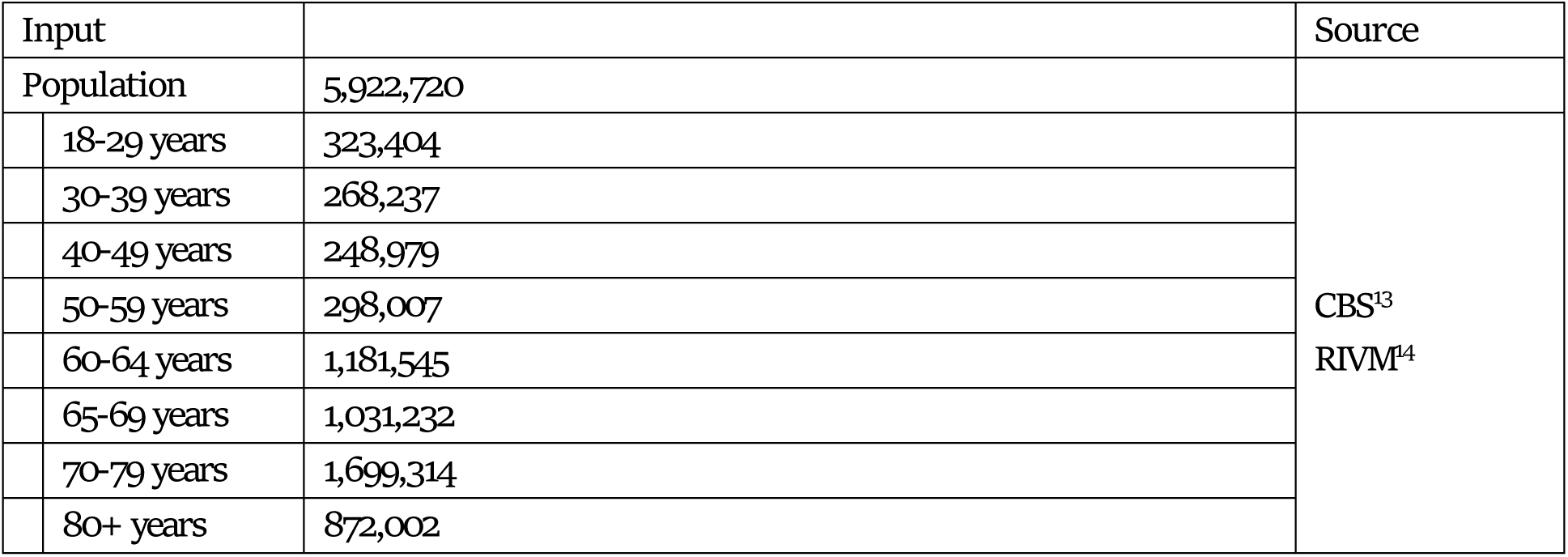
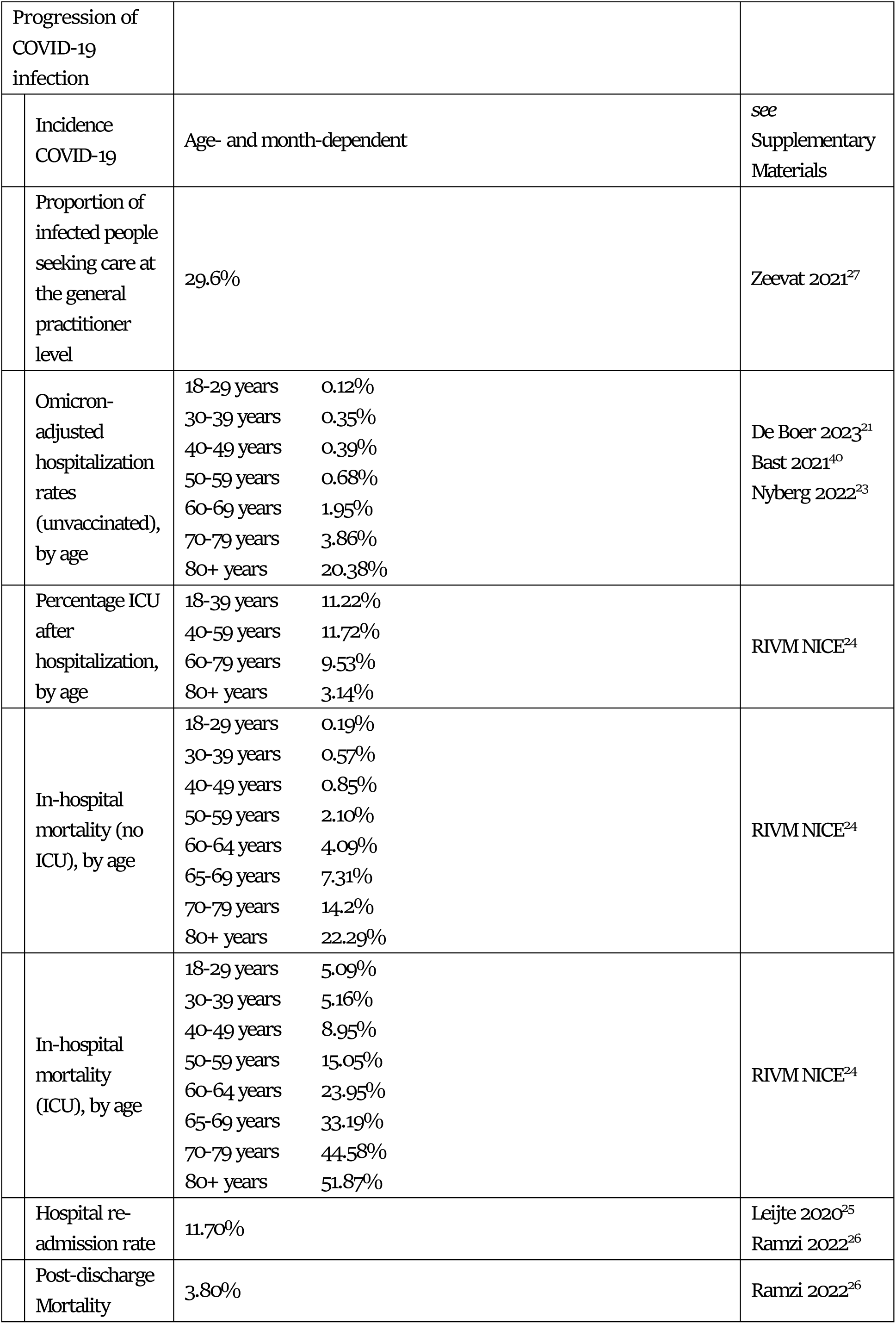
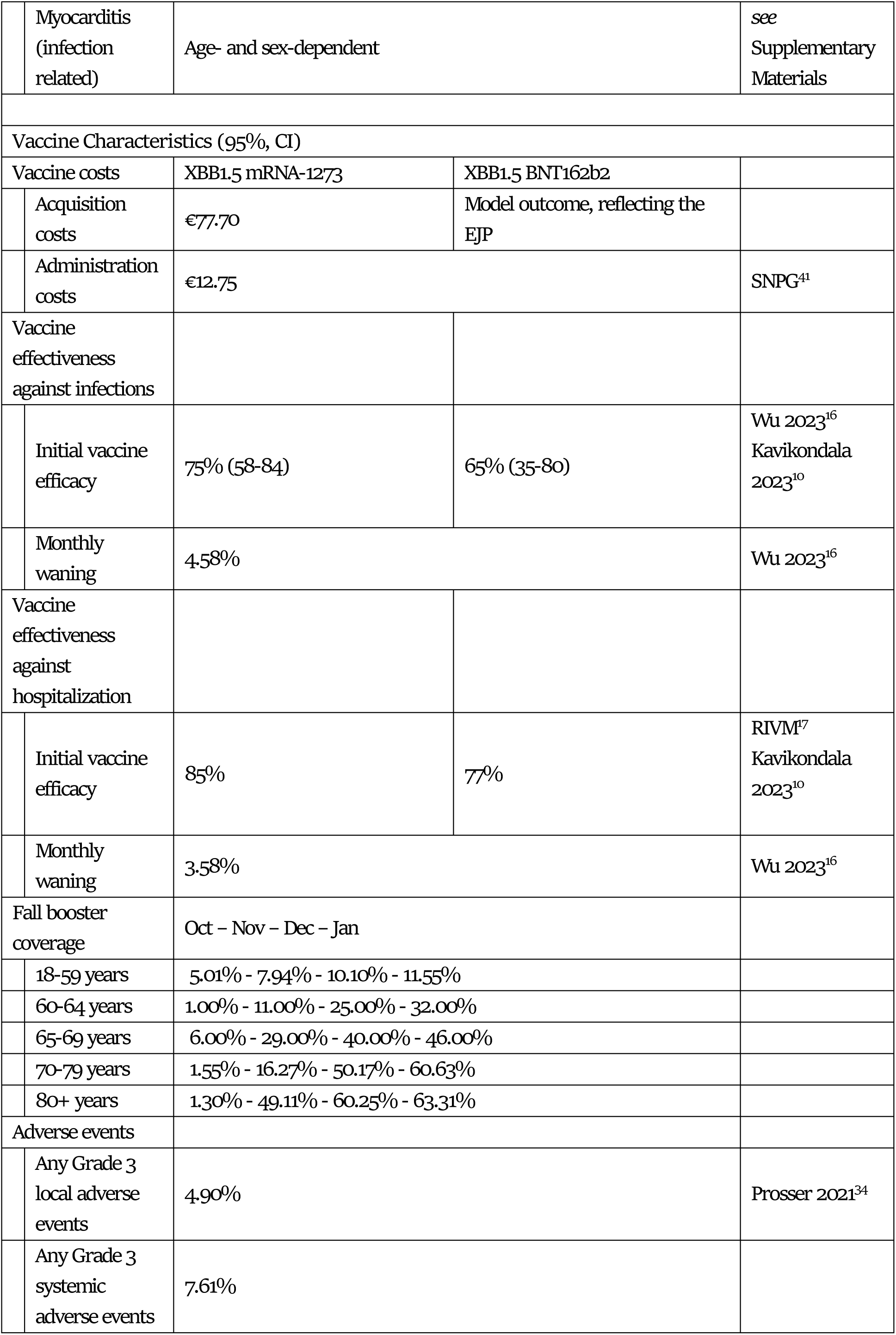
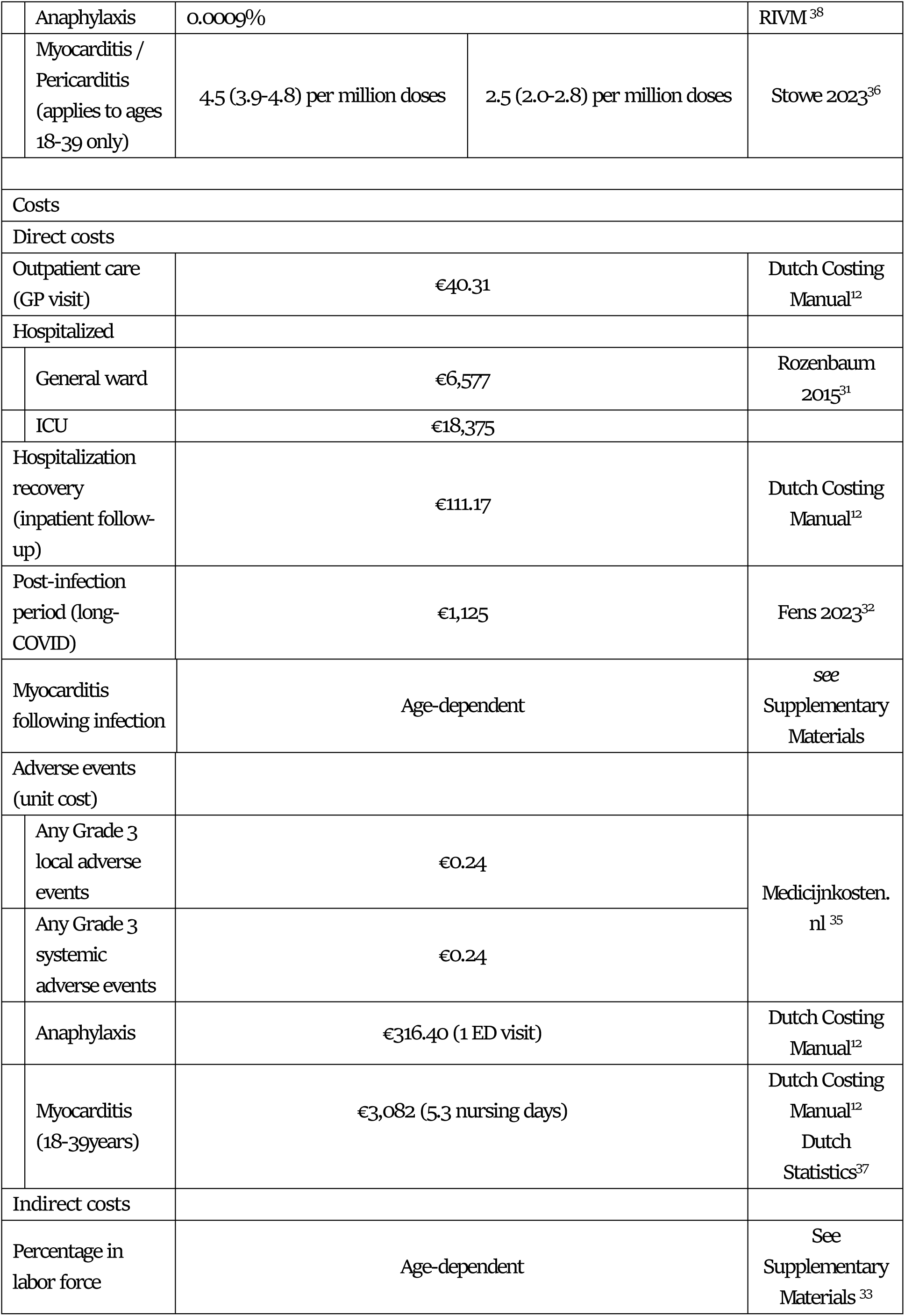
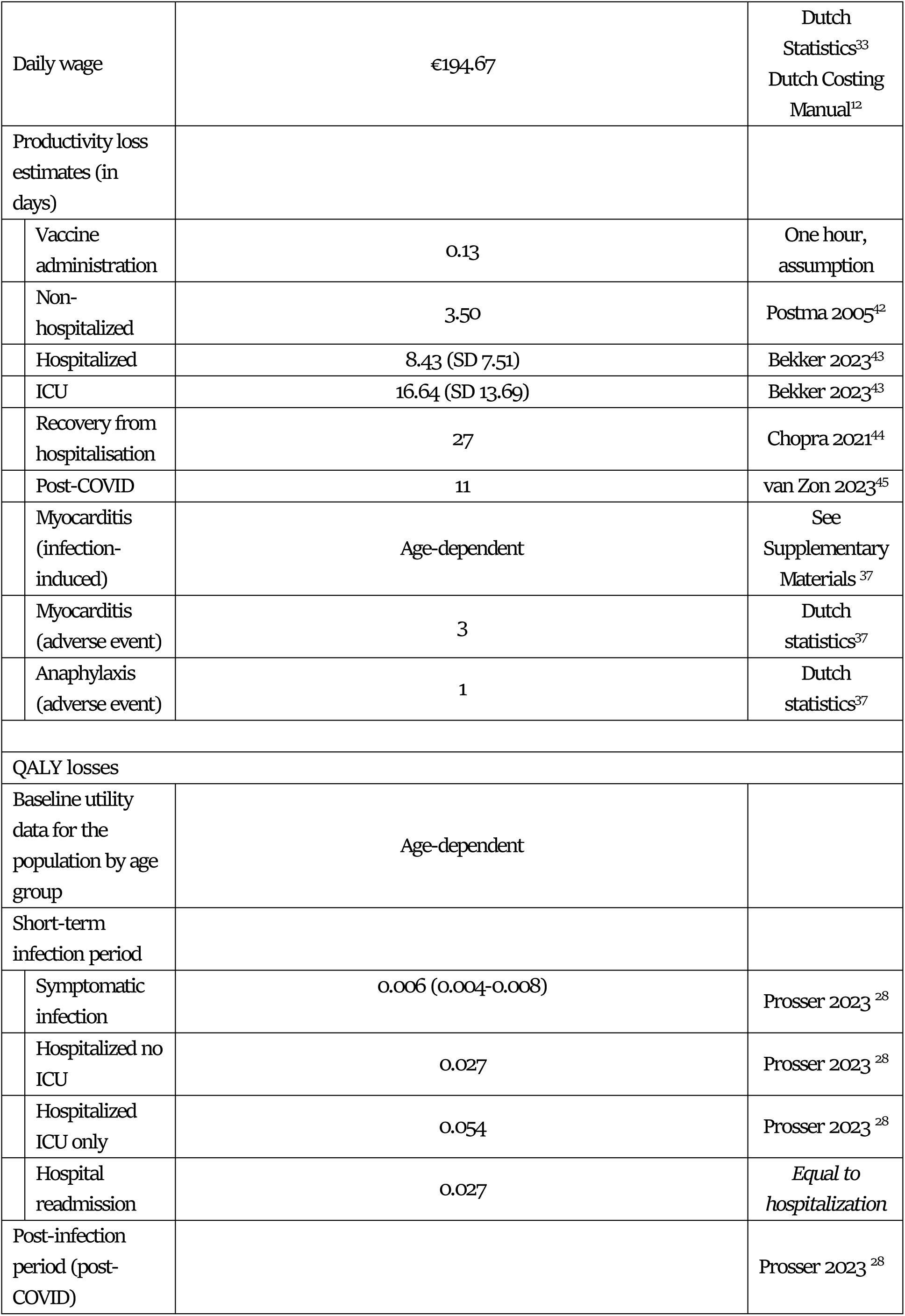
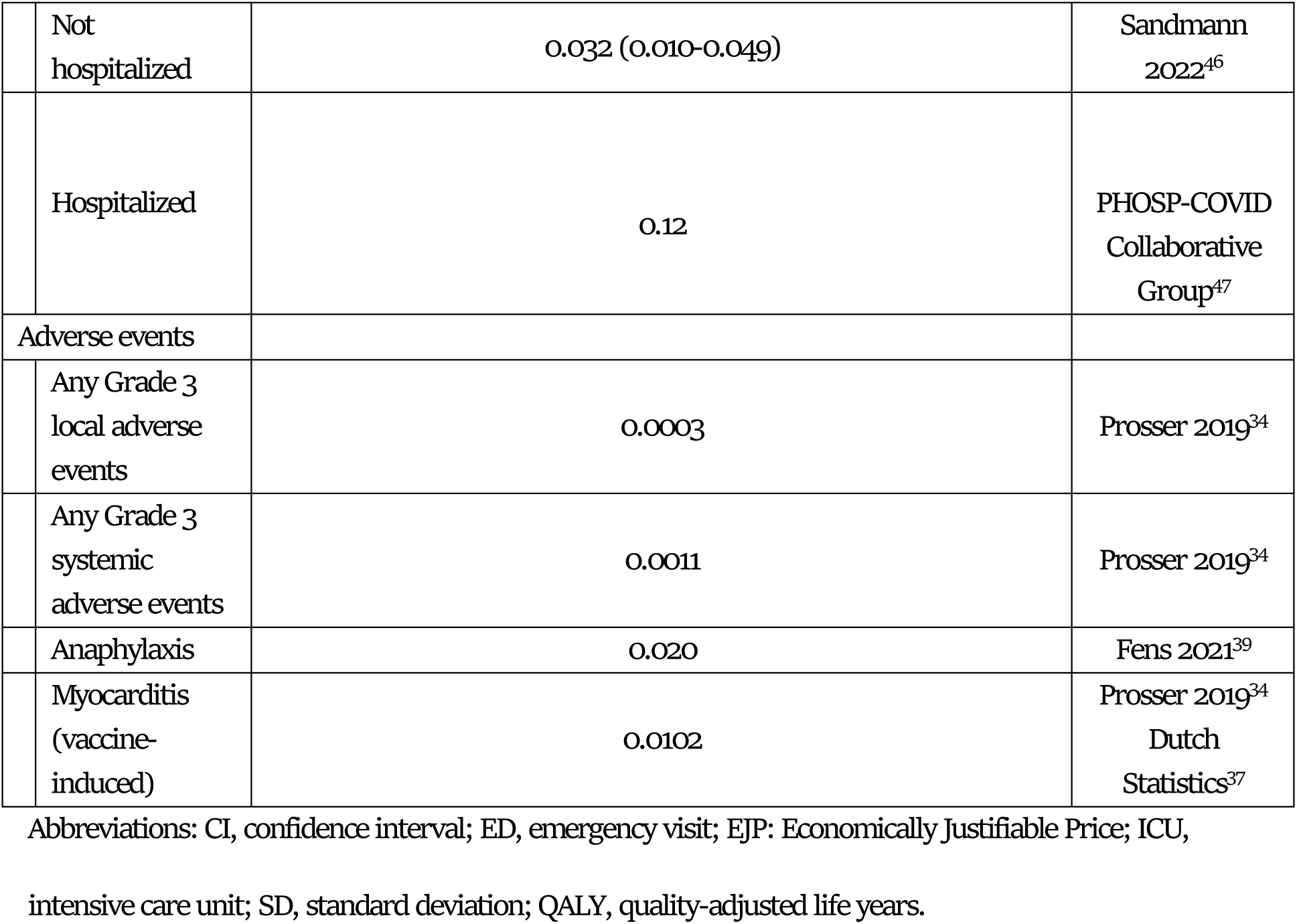
Input parameters.

### Target population

The population eligible for a booster vaccination was estimated at 5,922,719 (Table 1) and includes individuals aged 60 years and older, and individuals aged 18 to 59 years who are annually invited for influenza vaccination, including medical-risk groups, healthcare workers and pregnant individuals^8,13,14^.

### Vaccine characteristics

mRNA-1273 was compared with no Fall booster and with the BNT162b2 vaccine. The characteristics of both vaccines can be found in Table 1. The vaccine is administered from October to the end of December aligned with the Dutch recommendation. Consequently, vaccination coverage is obtained from the Fall vaccination campaign 2023^15^. For the older adults at risk (18-59 years), the number of vaccinated people in this age-group per month is divided by the estimated population and has a linear increase over the first two months, increasing from 0% to the coverage in the relevant age group on a monthly basis^8,13,14^

The vaccine effectiveness of BNT162b2 against symptomatic infection was derived from a meta-analysis showing a vaccine effectiveness of 65% at baseline against the omicron variant ^16^. For the effectiveness against hospitalisation, the risk reduction in hospital admissions in the Netherlands was applied at 77%^17^. The vaccine effectiveness of mRNA-1273 against symptomatic infection and hospitalization was assessed by using the effectiveness of BNT162b2 and the relative risk obtained from a meta-analysis^10,16,17^. This meta-analysis assessed the comparative effectiveness of mRNA-1273 versus BNT162b2 in the 50 years and older using 24 non-randomized real-world studies^10^, in which the relative risk for symptomatic infections and hospitalizations was 0.72 [95% CI 0.62‒0.83] and 0.65[95% CI 0.53‒0.79], respectively, favouring mRNA-1273. Additionally, the waning rate was determined by analyzing the BNT162b2 vaccine’s effectiveness at months 1, 4 and 5, and was considered equal for the mRNA-1273 vaccine^16^.

### Clinical input

The baseline risk of infection for COVID-19, or the incidence, was calculated using national hospitalization rates for COVID-19 during the 2022-2023 and 2023-2024seasons^18^, the vaccination coverage^19^, Dutch demographics^20^, and the unvaccinated hospitalization risk. More details can be found in the Supplementary Materials. Unvaccinated hospitalization risk was based on historical data from the pandemic^21^, corrected for the decreased severity of Omicron compared to the variants of the early pandemic^22,23^, see the Supplementary Materials for a detailed description of the calculation. The percentage ICU admissions following hospitalizations of the non-vaccinated per age-group was derived from The National Intensive Care Evaluation (NICE), a foundation that facilitates the registration of COVID-19 patients in Dutch ICU and general ward^24^. The data used covers the period from October 03, 2022, to May 29, 2023. Additionally, hospital-based mortality data, for both general ward and ICU admissions, were also derived from The National Intensive Care Evaluation ^24^. For hospital readmission and post-discharge mortality, only data from the pandemic period were available in the Netherlands, namely 11.70% and 3.80%, respectively^25,26^. For the proportion not hospitalized, influenza data were applied as no COVID-19 specific data were available^27^. In addition, following infection, myocarditis can occur depending on age. *See* the Supplementary Materials for estimates of the myocarditis/pericarditis age-dependent incidence, costs and QALY losses, including a detailed description of the calculation.

Neither in the Netherlands nor in Europe is data available on the health-related quality of life associated with symptomatic COVID-19 infections, including not-hospitalized cases, hospital admissions, ICU admissions, and post-covid conditions/long-COVID. As a result, our model incorporated US values^28^, QALY losses were derived from unpublished data obtained from the Coronavirus Household Evaluation and Respiratory Testing (C-HeaRT) and the Prospective Assessment of COVID-19 in a Community (PACC). Next to the QALY losses, life years lost were also included in the analysis.

### Costs

The actual acquisition price of the mRNA-1273 and BNT162b2 vaccine is unknown as these vaccines are purchased via European tenders and consequently prices are confidential. Nevertheless, the maximum price was assumed to be €77.70 based on the German listprice^29^. For BNT162b2, the Economically Justifiable Price (EJP) is estimated at a threshold value of 20,000 per QALY, which was the often-used willingness-to-pay (WTP) in the Fall of 2023 in the Netherlands. Early in 2024, are port was published where it was argued that the WTP threshold for public health interventions, including vaccines, should be increased to €50,000 per QALY^30^. This updated threshold was used in this analysis to assess the cost effectiveness of mRNA-1273.

Direct costs cover expenses associated with the treatment of COVID-19. In the Netherlands, hospital costs for both general ward and ICU care were not specified for COVID-19 but rather derived based on costs for individuals hospitalized with community-acquired pneumonia^31^. Additional healthcare costs were derived from the Dutch Costing manual^12^. Potential costs in the post-infection period (long-COVID) are €1125 in the first year in the Netherlands^32^, and only first-year costs are considered due to the limited availability of reliable long-term data and the variability in future healthcare needs. Productivity losses were included for the different health outcomes, as well as for adverse events, considering both the workforce participation rate and average daily wages^33^.

### Adverse events

For the adverse events related to the administration of the vaccines, we separated two main categories of adverse events. First, injection-related adverse events that are comparable to other vaccines and often are not severe and were extrapolated from data related to the herpes zoster vaccine. This dataset provides incidence rates and quality-adjusted life year (QALY) losses per adverse event (grade 3 systemic/local and severe events)^34^. For grade 3 adverse events following vaccination, expenses for painkillers were included^35^.

Second, for non-reactogenicity, and more severe adverse events, COVID-19 vaccine data were used. The occurrence of myocarditis was based on data from the United Kingdom and was specific for both COVID-19 vaccines included in the analysis^36^. For myocarditis, we assumed a QALY loss of 0.7, associated with a hospital stay duration of 5.3 days (based on the age group 20-45 years), resulting in a QALY loss per case of 0.0102 (calculated as 0.7/365*5.3)^34,37^. Myocarditis-related costs were also based on the hospital length of stay^12^. For anaphylaxis, Dutch COVID-19 specific incidence data were used^38^ and an influenza-related QALY loss was applied^39^. Costs per case were based on 1 emergency visit^12^.

### Analysis

The primary analysis considered the cost-effectiveness of mRNA-1273 compared to BNT162b2, expressed as an incremental cost-effectiveness ratio (ICER) as costs per QALY, from a societal perspective. For the primary analysis, the price BNT162b2 was estimated using a WTP of €20.000 per QALY compared to no vaccination, as explained above. Additionally, the price premium of mRNA-1273 compared to BNT162b2 was calculated using a WTP of €50.000 per QALY.

As stated above, tender prices are confidential in The Netherlands, therefore the mRNA-1273 was estimated assuming different BNT162b2 prices. In addition, to assess the robustness of the model, a deterministic sensitivity analysis was performed, quantifying the impact of specific parameters on the results. The distributions were varied by 20% of the mean. The 10most influential parameters were presented in a tornado diagram. In addition, a probabilistic sensitivity analysis was performed to evaluate the uncertainty in the results when all parameters were varied: involving 1,000 model runs with random samples from the underlying parameters distributions. These distributions in were based on the 95% Confidence Intervals (95%CI) or, in the absence of such intervals, varied by 20% from the mean. The results were presented in a cost-effectiveness plane and a cost-effectiveness acceptance curve (CEAC).

Next to sensitivity analyses, our study incorporated multiple scenario analyses comparing mRNA-1273 to BNT162b2, as detailed in Table 2. These analyses involved varying COVID-19 incidence rates, as the future incidence of COVID-19 is unknown as we transition from the pandemic to endemic phase, using both low and high estimated values, as explained in the Supplementary Materials. Moreover, one of the scenarios explored the incidence of myocarditis following infection and vaccination, utilizing data from the United States, where no difference between the vaccines was observed^48^.

**Table 2.**
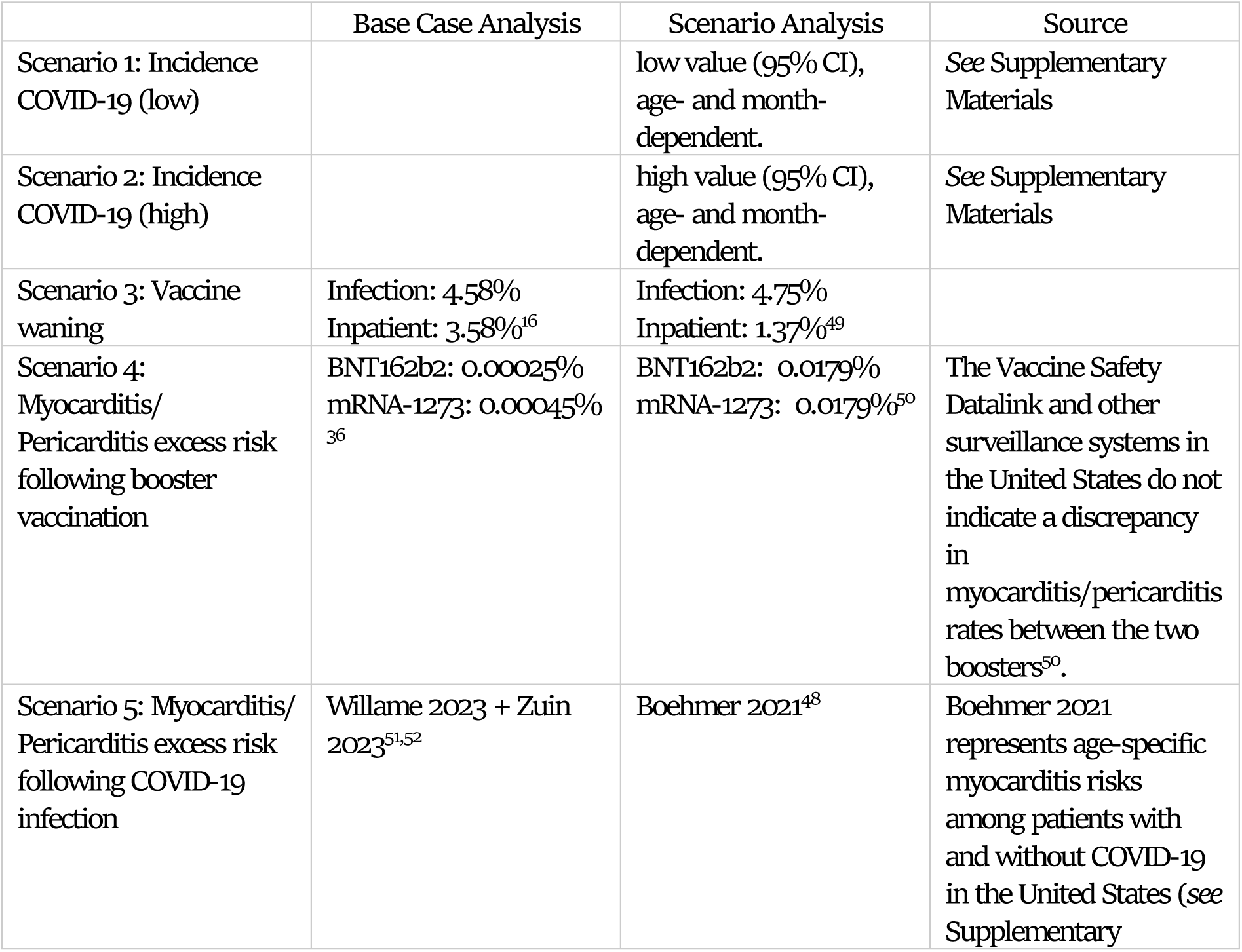
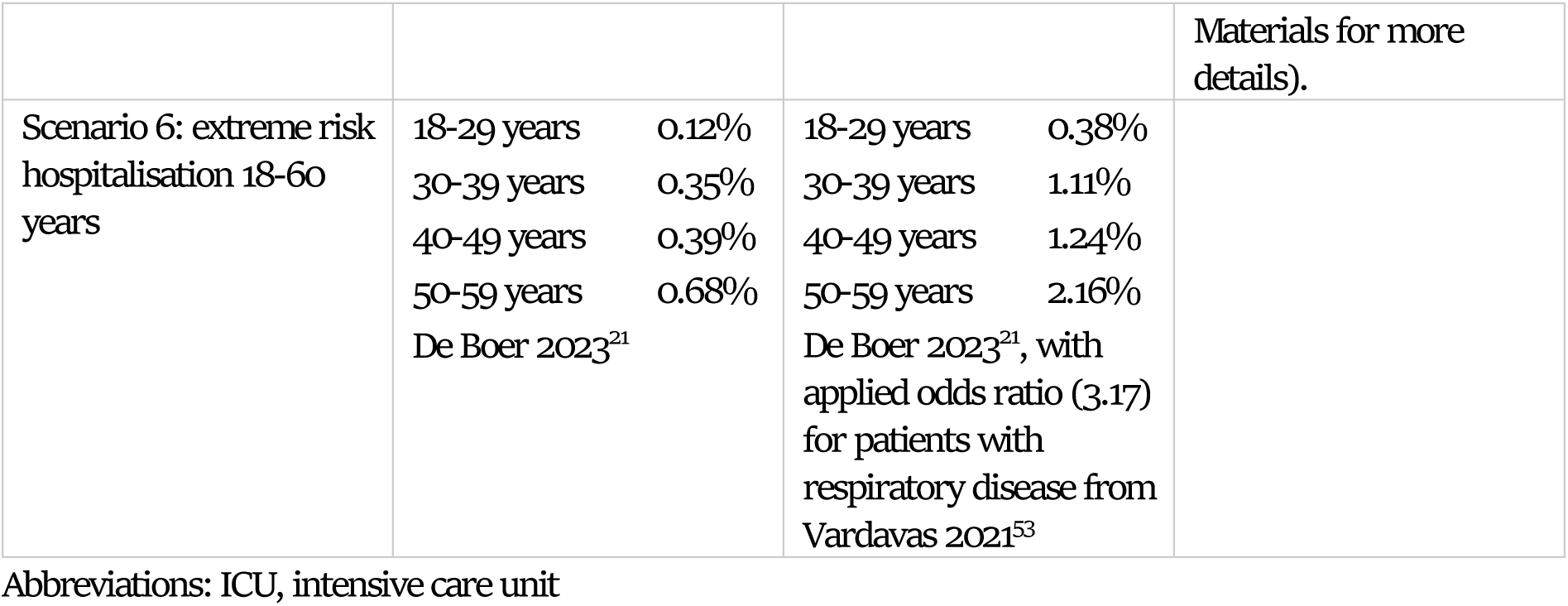
Scenario analysis.

## Results

### Base case

The public COVID vaccination program targeting 5,922,720 people of which approximately 2,566,389 people are being vaccinated substantially reduces the burden of COVID disease. In particular, it was estimated that 110,604 symptomatic infections, 2667 hospitalisations and 604 deaths are averted. The EJP for BNT126b2 was determined compared to no vaccination, based on a WTP of €20,000 or €50,000 per QALY, resulted in a price of €38.41 or €92.97 per dose, respectively.

When comparing BNT126b2 to mRNA-1273, for the high-risk population, 20,629 symptomatic cases can be averted including 924 hospitalizations of which 32 ICU admissions. A total of 207 deaths can be averted.

Excluding the potential additional investment of mRNA-1273, significant reductions in costs are expected, with almost an additional €10million saved from the healthcare perspective and €2.9 million productivity gained in the population. The health gains relate to 1,506QALYs gained (Table 3), see Supplementary Materials for more detailed results. While mRNA-1273 is linked to on average 0.12 more vaccination-related myocarditis cases, 26 more infection-related myocarditis cases can be prevented compared to BNT126b2.

**Table 3:**
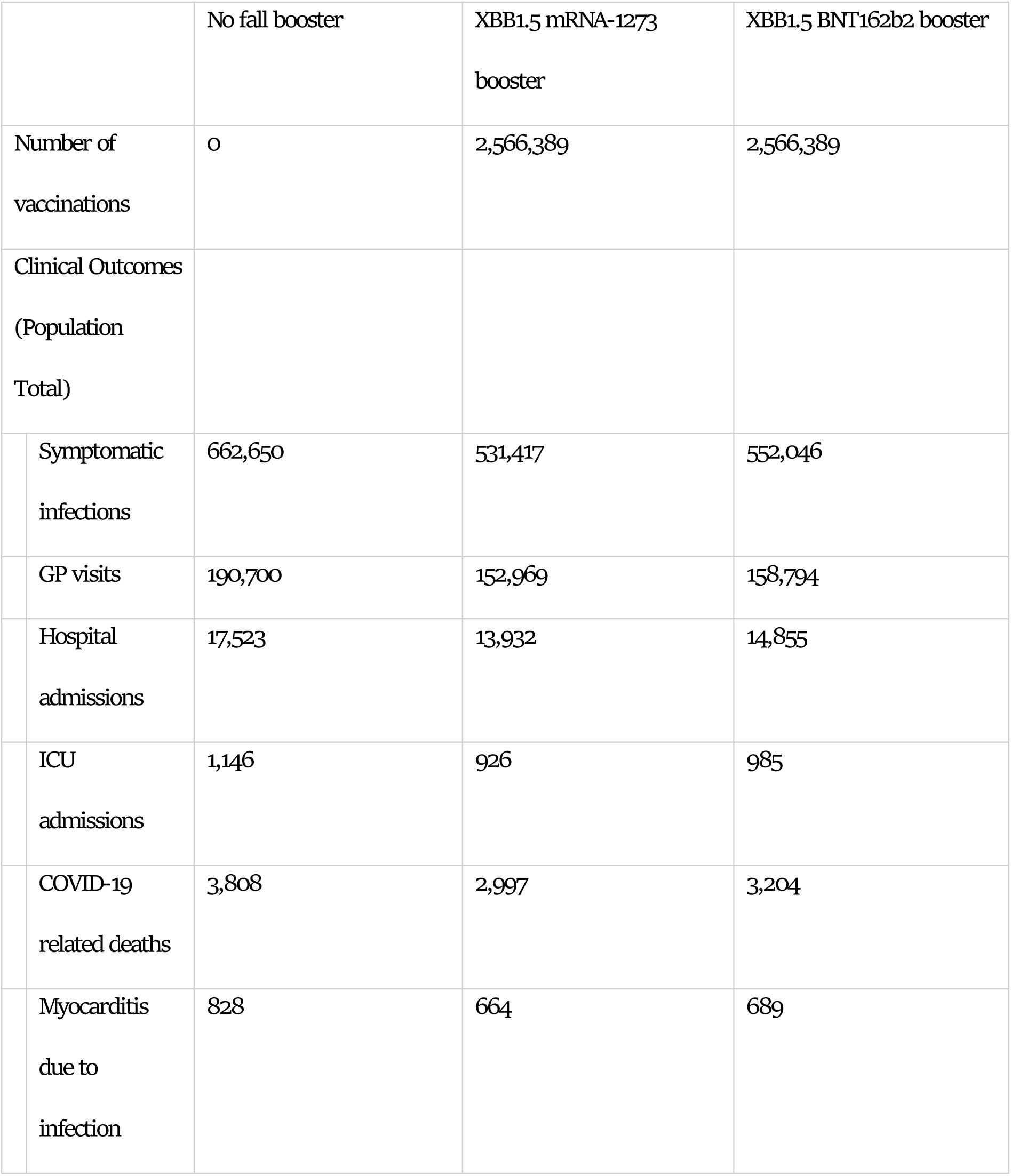
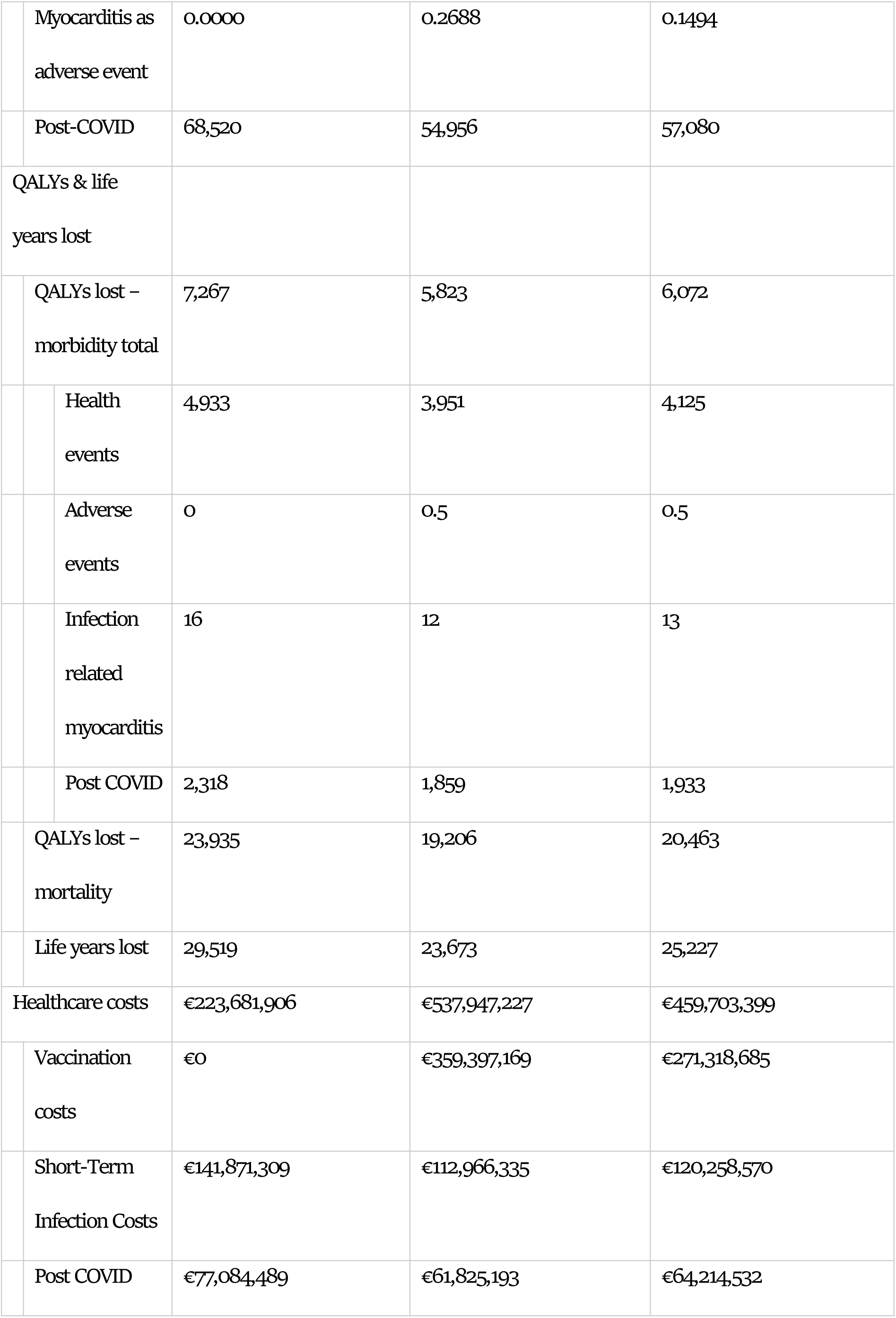
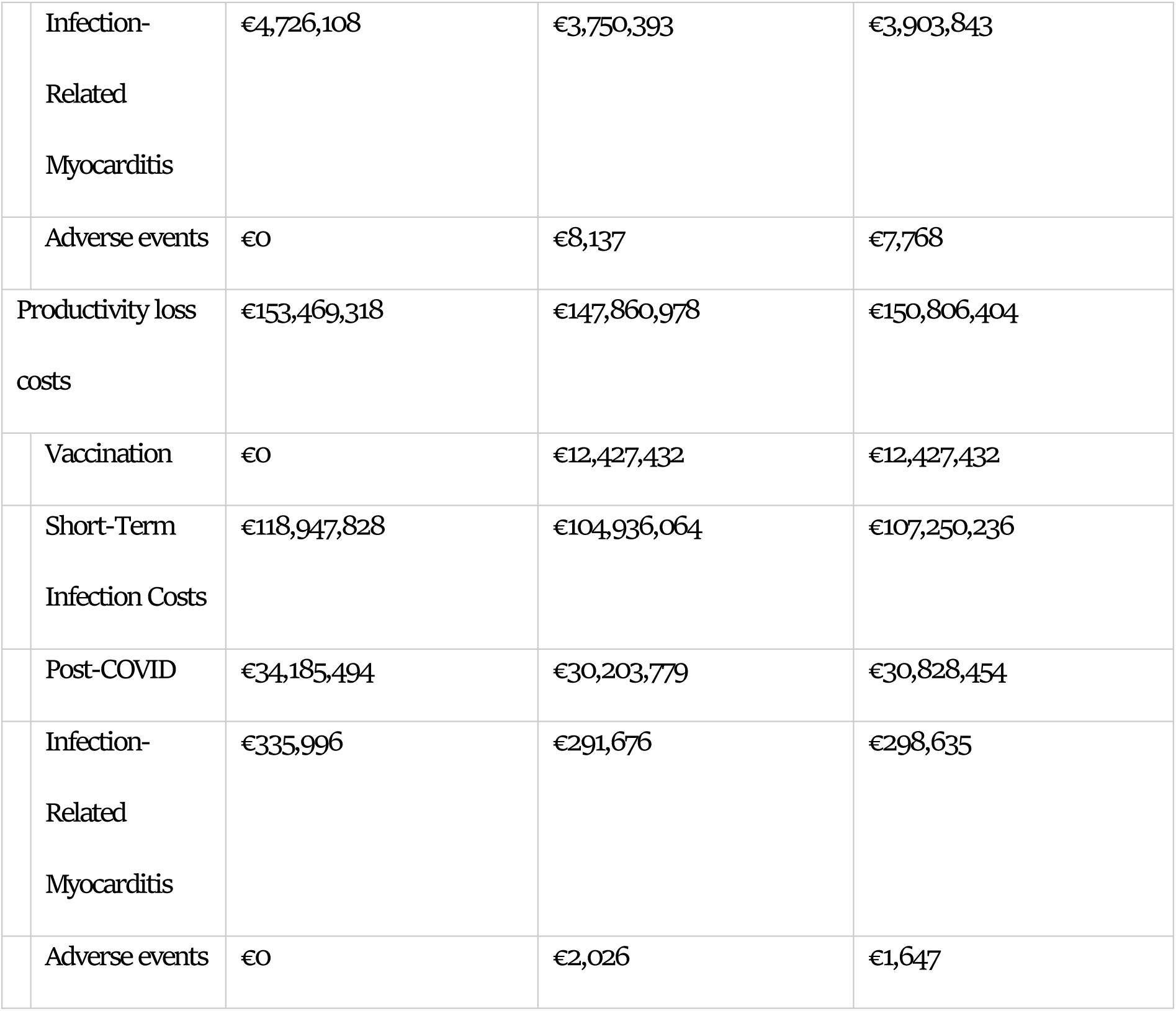
Health and cost outcomes model.

|  | No fall booster | XBB1.5 mRNA-1273<br>booster | XBB1.5 BNT162b2 booster |
| --- | --- | --- | --- |
| Number of<br>vaccinations | 0 | 2,566,389 | 2,566,389 |
| Clinical Outcomes<br>(Population<br>Total) |  |  |  |
| Symptomatic<br>infections | 662,650 | 531,417 | 552,046 |
| GP visits | 190,700 | 152,969 | 158,794 |
| Hospital<br>admissions | 17,523 | 13,932 | 14,855 |
| ICU<br>admissions | 1,146 | 926 | 985 |
| COVID-19<br>related deaths | 3,808 | 2,997 | 3,204 |
| Myocarditis<br>due to<br>infection | 828 | 664 | 689 |

|  |  |  |  |  |
| --- | --- | --- | --- | --- |
|  | Myocarditis as adverse event | 0.0000 | 0.2688 | 0.1494 |
|  | Post-COVID | 68,520 | 54,956 | 57,080 |
|  | QALYs & life years lost |  |  |  |
|  | QALYs lost – morbidity total | 7,267 | 5,823 | 6,072 |
|  | Health events | 4,933 | 3,951 | 4,125 |
|  | Adverse events | 0 | 0.5 | 0.5 |
|  | Infection related myocarditis | 16 | 12 | 13 |
|  | Post COVID | 2,318 | 1,859 | 1,933 |
|  | QALYs lost – mortality | 23,935 | 19,206 | 20,463 |
|  | Life years lost | 29,519 | 23,673 | 25,227 |
|  | Healthcare costs | €223,681,906 | €537,947,227 | €459,703,399 |
|  | Vaccination costs | €0 | €359,397,169 | €271,318,685 |
|  | Short-Term Infection Costs | €141,871,309 | €112,966,335 | €120,258,570 |
|  | Post COVID | €77,084,489 | €61,825,193 | €64,214,532 |
| Infection-Related<br>Myocarditis | €4,726,108 | €3,750,393 | €3,903,843 |
| Adverse events | €0 | €8,137 | €7,768 |
| Productivity loss<br>costs | €153,469,318 | €147,860,978 | €150,806,404 |
| Vaccination | €0 | €12,427,432 | €12,427,432 |
| Short-Term<br>Infection Costs | €118,947,828 | €104,936,064 | €107,250,236 |
| Post-COVID | €34,185,494 | €30,203,779 | €30,828,454 |
| Infection-Related<br>Myocarditis | €335,996 | €291,676 | €298,635 |
| Adverse events | €0 | €2,026 | €1,647 |

Both vaccination strategies are effective in preventing health effects and costs when compared to no fall booster vaccination in the Netherlands (Table 3). Considering WTP thresholds of €20,000 and €50,000 per QALY gained, the maximum price premium of mRNA-1273 compared to BNT126b2 should be €16.72 or €34.32 per dose, resulting in total prices per dose of €55.12 and €127.29, respectively.

### Sensitivity analyses

Figure 2 shows the tornado diagram of the deterministic sensitivity analysis. The most influential parameters are the vaccination costs, followed by the initial vaccine effectiveness, hospitalization rate, and baseline utility values. Figure 2A shows the outcomes assuming a WTP threshold of €20,000 and 2B a WTP of €50,000 per QALY gained.

**Figure 2.**
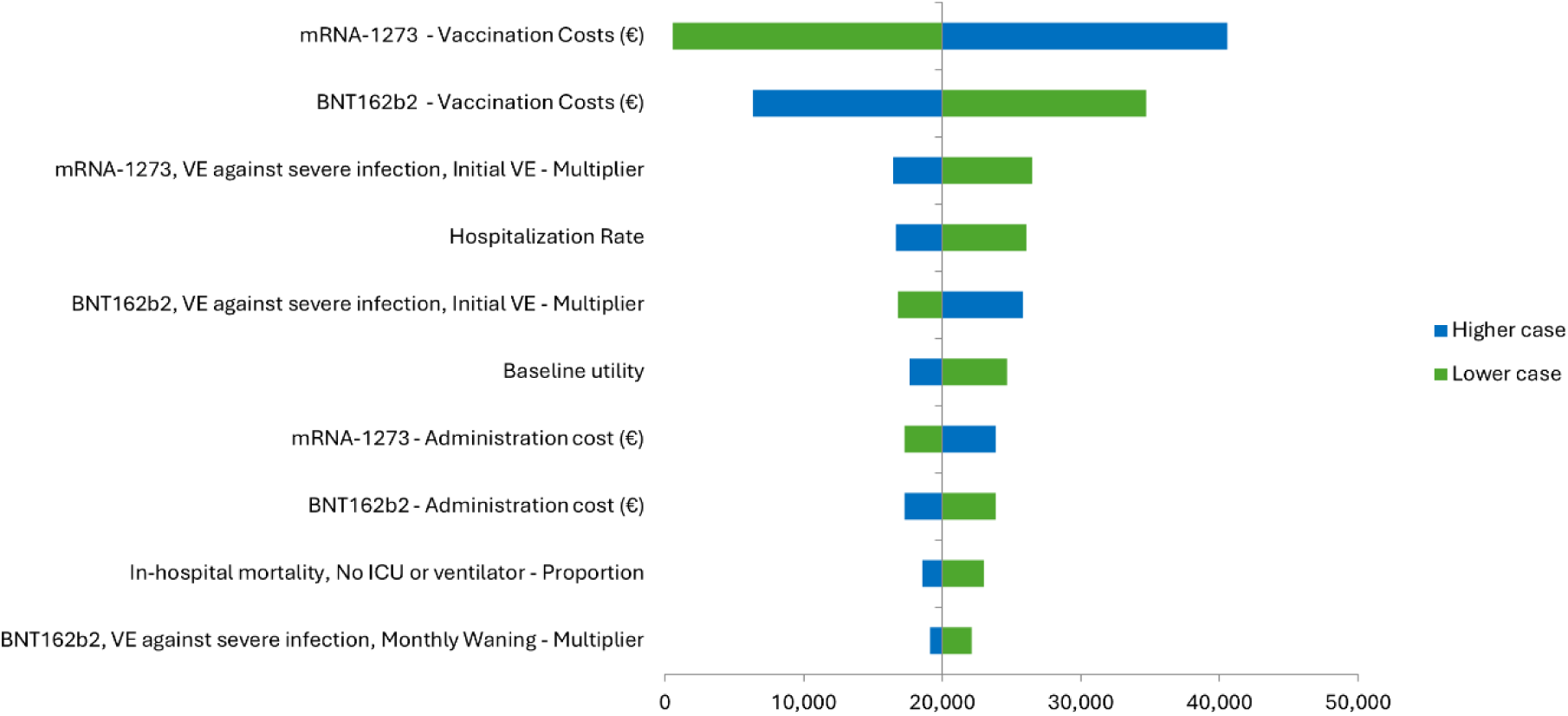
Tornado plot of the deterministic sensitivity analysis, showing the 10 most influential parameters in the model The CEAC (Figure 3) shows that mRNA-1273 is approximately 40% likely to be cost-effective at the threshold of €20,000 per QALY gained considering a €16,72 price difference. Considering a threshold of €50,000/QALY 95% of simulations were found to consider mRNA-1273 cost-effective. The CE plane is included in the appendix.

**Figure 3.**
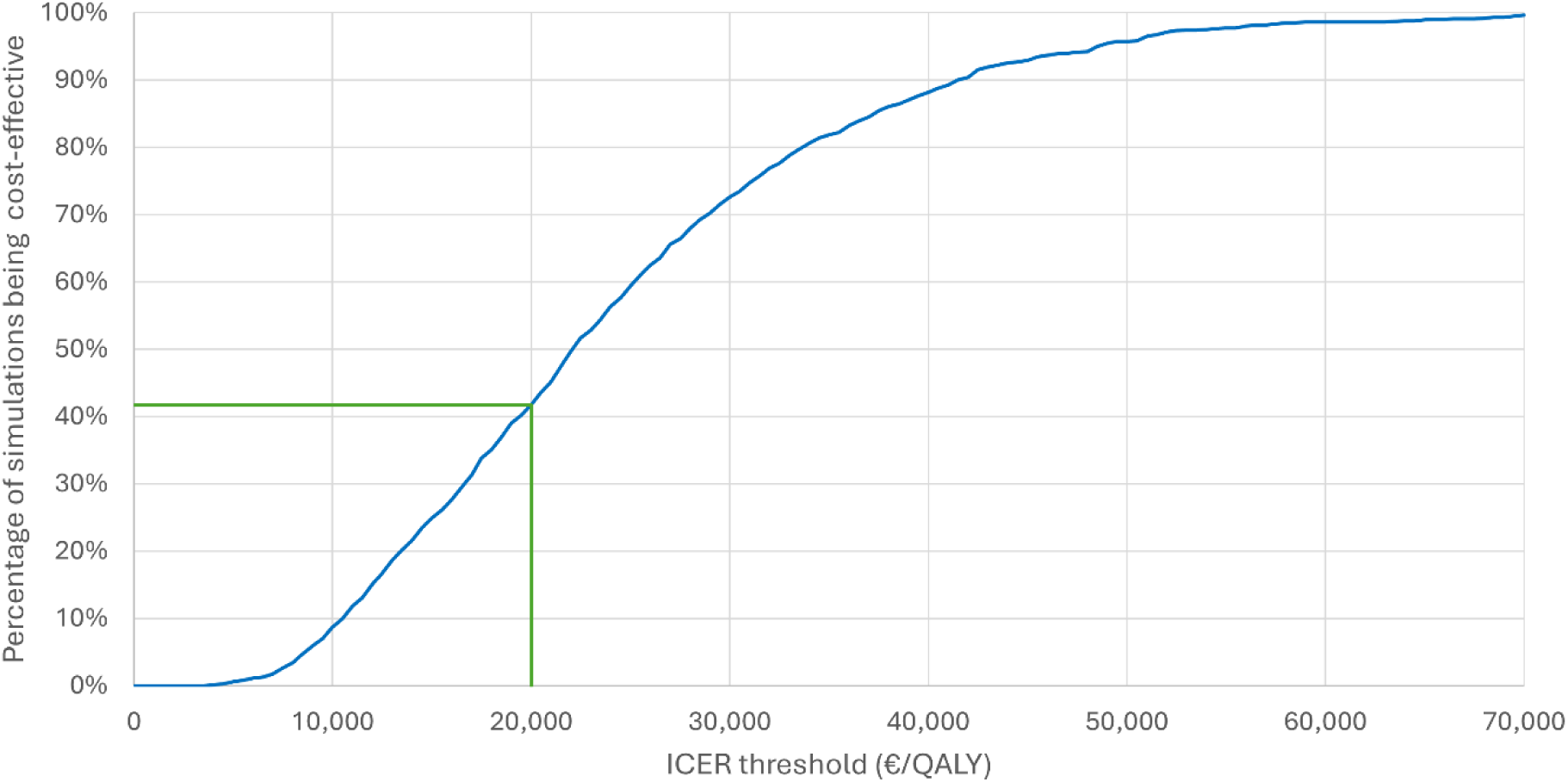
Cost-Effectiveness Acceptability Curve of a nation-wide vaccination programme for the elderly and risk groups with XBB1.5 mRNA-1273 compared to XBB1.5 BNT126b2 for the Netherlands. Al costs are displayed in Euros. ICER: Incremental Cost-Effectiveness Ratio.

### Scenario analyses

The scenario analyses are included in Table 4. The scenarios with the low and high incidence have the most impact on the ICER, ranging from €28,718 per QALY under a high incidence scenario to €96,439 per QALY when the incidence is low. The impact from other scenarios on the ICER were negligible.

**Table 4.** scenario analyses comparing XBB1.5 mRNA-1273 to XBB1.5 BNT126b2.

|  | Costs (societal perspective) | Effects (QALYs) | ICER |
| --- | --- | --- | --- |
| Base case | XBB1.5 mRNA-1273:<br>€500,591,883<br>XBB1.5 BNT126b2:<br>€470,492,350<br>Incremental:<br>€30,099,533 | XBB1.5 mRNA-1273:<br>25,028<br>XBB1.5 BNT126b2:<br>26,534<br>Incremental: 1,506 | €19,989 per QALY |
| Scenario 1: Incidence COVID-19 (low) | XBB1.5 mRNA-1273:<br>€398,556,049<br>XBB1.5 BNT126b2:<br>€363,886,720<br>Incremental:<br>€34,670,329 | XBB1.5 mRNA-1273:<br>16,274<br>XBB1.5 BNT126b2:<br>17,234<br>Incremental: 960 | €36,100 per QALY |
| Scenario 2: Incidence COVID-19 (high) | XBB1.5 mRNA-1273:<br>€701,015,632<br>XBB1.5 BNT126b2:<br>€680,946,352<br>Incremental:<br>€20,069,280 | XBB1.5 mRNA-1273:<br>43,725<br>XBB1.5 BNT126b2:<br>46,442<br>Incremental: 2,717 | €7387 per QALY |
| Scenario 3: Vaccine waning | XBB1.5 mRNA-1273:<br>€495,378,409<br>XBB1.5 BNT126b2:<br>€465,278,877<br>Incremental:<br>€30,099,533 | XBB1.5 mRNA-1273:<br>24,060<br>XBB1.5 BNT126b2:<br>25,565<br>Incremental: 1,506 | €19,989 per QALY |
| Scenario 4: Myocarditis/ Pericarditis excess risk following booster vaccination | XBB1.5 mRNA-1273:<br>€500,657,069<br>XBB1.5 BNT126b2:<br>€470,558,283<br>Incremental:<br>€30,098,786 | XBB1.5 mRNA-1273:<br>25,028<br>XBB1.5 BNT126b2:<br>26,534<br>Incremental: 1,506 | €19,988 per QALY |
| Scenario 5: Myocarditis/ Pericarditis excess risk following COVID-19 infection | XBB1.5 mRNA-1273:<br>€501,834,999<br>XBB1.5 BNT126b2:<br>€471,798,737<br>Incremental:<br>€30,036,262 | XBB1.5 mRNA-1273:<br>25,032<br>XBB1.5 BNT126b2:<br>26,538<br>Incremental: 1,506 | €19,944 per QALY |
| Scenario 6: extreme risk hospitalisation 18-60 years | XBB1.5 mRNA-1273:<br>€508,603,585<br>XBB1.5 BNT126b2:<br>€478,630,618<br>Incremental:<br>€29,972,967 | XBB1.5 mRNA-1273:<br>25,762<br>XBB1.5 BNT126b2:<br>27,275<br>Incremental: 1,513 | €19,812 per QALY |

## Discussion

In current study, the objective was to conduct a comprehensive cost-effectiveness analysis of a COVID-19 vaccination campaign in the Fall of 2023 with mRNA-1273, compared to BNT162b2, in the high-risk population, in the context of the Netherlands. The urgency of this research stems from the need to inform public health decision-makers about the potential benefits and economic implications associated with the adoption of either vaccine in high-risk groups. Both vaccines provide major population health benefits for the health of the population. Our findings show that the XBB.1.5 mRNA-1273 vaccine, is projected to provide substantial additional clinical benefits compared to BNT162b2. Specifically, it is estimated that it can prevent an additional 20,629 symptomatic cases, 924 hospital admissions (including 32 ICU admissions), and 207 deaths compared to BNT162b2. These health effects translate to €10 million healthcare costs averted(without vaccination costs), €2.9 million in productivity gains, and 1,506 total QALYs gained.

For this analysis the price of BNT162b2 was calculated using the EJP and a WTP of €20.000 and €50,000 per QALY. Vaccine prices commonly are subject to tenders, making the comparison of list prices challenging. Confidential discounts on list prices are provided by vaccine manufacturers in tenders, subject to other terms such as predictable supply, long-term agreements and in some cases market exclusivity. Considering the currently available list price of mRNA-1273 of €77.70, COVID-19 vaccination can be perceived as cost-effective against a WTP of €50,000 per QALY as we estimated the EJP of mRNA-1273 at €55.12 or €127.29 per dose considering a WTP of €20,000 and €50,000 per QALY, respectively. The EJP of BNT162b2 was estimated at €38.41 or €92.97 per dose, considering both WTP thresholds.

One of the identified risks associated with booster vaccination is the potential occurrence of myocarditis or pericarditis. There is a higher observed risk of myocarditis and/or pericarditis in the population aged 18-39 years, particularly in males aged 18-29 years^36,54–57^. Specifically, the mRNA-1273 vaccine has been associated with a slightly higher risk of myocarditis and/or pericarditis in comparison to the BNT162b2 vaccine within this population^58^. However, myocarditis risk after booster vaccination is significantly lower compared to the primary series regimen of 2 x100µg^36,57^. The high-risk group for myocarditis-as a side effect and complication of infection - i.e. males aged 18-39 with comorbidities, account for a small proportion of the target group for booster vaccination, thus the estimated difference in the number of cases of myocarditis between mRNA-1273 and BNT162b2 is relatively small in current analysis (0.2688 cases with mRNA-1273 vs 0.1494 cases with BNT162b2). When considering mRNA COVID-19 vaccination risk-benefit with regards to myocarditis, the relative risk for myocarditis is more than seven times higher in patients with COVID-19 compared than those vaccinated^59^, resulting in more infection-related myocarditis averted with the mRNA-1273 booster compared toBNT162b2(664caseswithmRNA-1273vs689cases with BNT162b2).Considering the public health benefits of vaccination programmes against COVID-19, mRNA COVID-19 vaccines have a positive risk-benefit profile.

This is the first cost-effectiveness analysis that estimates the cost-effectiveness of the two vaccines for a Fall vaccination campaign in the Netherlands. Two similar studies have been published for other countries, one for Germany and one for the United States^49,60^. Both studies used a dynamic modelling approach to estimate the incidence of COVID-19. For Germany, a price premium based on the EJP of €24.95 per dose was found for mRNA-1273 at a WTP threshold of €50.000 per QALY^60^. This is highly comparable to our results, which yielded a premium of €34.32 for mRNA-1273 at the same threshold. For the United States, vaccine prices of $129.59 and $120.00 were used for mRNA-1273 and BNT162b2, respectively, yielding an ICER of $2,100 per QALY from the societal perspective, which is also aligned with our results.

There are some limitations to the present study. First, the challenge of accurately capturing the incidence of COVID-19 poses an important limitation. Therefore, we performed a scenario analysis with lower and higher incidence values than assumed, where we showed that the incidence is a key influential parameter. This stresses the importance for health systems worldwide to implement adequate surveillance systems on pathogens circulating within the populations; it is currently not possible to obtain accurate incidence numbers for COVID-19 in the Netherlands, other than hospital admissions. For the base case analysis, we can compare the number of hospitalizations in the model to the actual reported number of hospitalizations.

In the model 14,855 hospitalizations are expected in the XBB1.5 BNT162b2 booster arm, while in the year prior to mid-February 2024, a total of 15,238 hospitalizations have been reported for the Netherlands^18^. A second limitation was the absence of head-to-head randomized clinical trials, comparing the vaccine effectiveness of mRNA-1273 and the BNT162b2. Instead, we relied on risk estimates derived from a recent meta-analysis^10^. In our analysis, we did not account for any potential wastage of either vaccine resulting from the different presentations, i.e. mRNA-1273 as prefilled syringe and BNT162b2 as either multi-dose or single-dose vial. Furthermore, the hospital costs in our study were based on a specific study focusing on hospital admissions with community-acquired pneumonia^31^. This choice may not fully account for the potentially higher costs associated with COVID-19 hospitalizations, such as prolonged hospital stays and the use of mechanical ventilation. The duration of hospitalization nowadays, particularly with the new variants, might be shorter compared to the early phases of the pandemic. Regarding the hospitalization rate, estimates from the general population were used^21^, not accounting for the high-risk group targeted in this analysis. Applying an extreme increased risk, based on all targeted patients having respiratory disease^53^, proved to have a marginal impact on the ICER. Lastly, our model exclusively includes vaccination-induced myocarditis for the age group of 18-40 years, despite its occurrence in other age groups^36^. Despite this limitation, our scenario and sensitivity analysis on the incidence of vaccine-induced myocarditis showed a negligible effect on the overall outcome.

While the mRNA-1273is more effective compared to the BNT162b2 vaccine, the Dutch government exclusively purchased the BNT162b2. This suggests missed opportunities for optimizing public health outcomes; our analysis reveals that utilizing mRNA-1273 instead of BNT162b2 could prevent a significant number of additional symptomatic cases as well as reduce hospitalizations and ICU admissions. Therefore, future vaccination recommendations in the Netherlands should consider a more inclusive approach that evaluates and incorporates the comparative (cost-)effectiveness of available vaccine options. This strategy aims to maximize health benefits for the population.

The cost-effectiveness of mRNA-1273 is dependent on the price of its competition (BNT126b2), and while it is uncertain whether it is cost-effective at its list price, it is likely to be cost-effective as long as the Dutch government negotiates a maximum price premium of €34.32 compared to BNT162b2.

## Data Availability

All data produced in the present work are contained in the manuscript

