## Supplementary Materials for "Cost-effectiveness analysis of COVID-19 mRNA XBB.1.5 Fall 2023 vaccination in the Netherlands"

### Methods

##### *Calculation of unvaccinated hospitalization risk*

The hospitalization risk for the unvaccinated population was calculated based on data before the initial COVID-19 vaccine rollout during the pandemic, corrected for the delta and omicron variants of the virus. In the table below, this can be calculated as: a x b x c.

| **Age group** | **Risk COVID hospitalization**[1] *a* | **Relative risk pre-delta vs. delta variant (95% CI)**[2] *b* | **Relative risk omicron vs. delta variant**[3] *c* | **Calculated risk hospitalization unvaccinated** |
|---|---|---|---|---|
| 20-29 years | 0.14% | 1.93 (1.84 – 2.03) | 0.43 (0.37 – 0.49) | 0.12% |
| 30-39 years | 0.59% | | 0.31 (0.28 – 0.35) | 0.35% |
| 40-49 years | 1.00% | | 0.20 (0.17 – 0.23) | 0.39% |
| 50-59 years | 2.50% | | 0.14 (0.12 – 0.17) | 0.68% |
| 60-69 years | 7.20% | | 0.14 (0.12 – 0.16) | 1.95% |
| 70-79 years | 10.0% | | 0.20 (0.17 – 0.24) | 3.86% |
| 80+ years | 32.0% | | 0.33 (0.28 – 0.39) | 20.38% |

##### *Calculation of risk of infection*

The risk of infection was calculated using the hospitalization risk for the unvaccinated (see above), the number of hospitalization in the period September 2022 to July 2023[4], the vaccination coverage in both the general population and the hospitalized population[5], and statistics on the size of the general population[6].

The results are displayed on the following page.

| | Hospitalizations | Vaccination coverage | Unvaccinated hospitalizations[1] | Calculated risk of hospitalization[1] | Backcalculated number of cases[1] | Unvaccinated population[1] | Risk of COVID-19 infection[1,2] |
|---|---|---|---|---|---|---|---|
| January | | | | | | | |
| 20-29 years | 25 | 10% | 22 | 0.12% | 19,366 | 2,233,707 | 0.87% (0.60% - 1.25% |
| 30-39 years | 49 | 10% | 44 | 0.35% | 12,493 | 2,171,853 | 0.58% (0.39% - 0.82% |
| 40-49 years | 60 | 17% | 50 | 0.39% | 12,902 | 2,157,398 | 0.60% (0.43% - 0.82% |
| 50-59 years | 161 | 17% | 134 | 0.68% | 19,782 | 2,440,051 | 0.81% (0.60% - 1.09% |
| 60-64 years | 130 | 42% | 75 | 1.95% | 3,876 | 639,365 | 0.61% (0.43% - 0.83% |
| 65-69 years | 219 | 42% | 127 | 1.95% | 6,529 | 564,123 | 1.16% (0.83% - 1.59% |
| 70-79 years | 670 | 42% | 389 | 3.86% | 10,067 | 523,291 | 1.92% (1.30% - 2.91% |
| 80+ years | 898 | 53% | 422 | 20.38% | 2,071 | 275,081 | 0.75% (0.35% - 1.77% |
| February | | | | | | | |
| 20-29 years | 40 | 1% | 40 | 0.12% | 34,083 | 2,233,707 | 1.53% (1.06% - 2.19% |
| 30-39 years | 62 | 1% | 61 | 0.35% | 17,388 | 2,171,853 | 0.80% (0.54% - 1.15% |
| 40-49 years | 91 | 25% | 68 | 0.39% | 17,681 | 2,157,398 | 0.82% (0.60% - 1.14% |
| 50-59 years | 173 | 25% | 130 | 0.68% | 19,208 | 2,440,051 | 0.79% (0.59% - 1.07% |
| 60-64 years | 167 | 48% | 87 | 1.95% | 4,464 | 639,365 | 0.70% (0.50% - 0.96% |
| 65-69 years | 244 | 48% | 127 | 1.95% | 6,522 | 564,123 | 1.16% (0.83% - 1.59% |
| 70-79 years | 814 | 48% | 423 | 3.86% | 10,966 | 523,291 | 2.10% (1.42% - 3.17% |
| 80+ years | 1084 | 57% | 466 | 20.38% | 2,287 | 275,081 | 0.83% (0.38% - 1.95% |
| March | | | | | | | |
| 20-29 years | 36 | 13% | 31 | 0.12% | 26,957 | 2,233,707 | 1.21% (0.84% - 1.73% |
| 30-39 years | 67 | 13% | 58 | 0.35% | 16,513 | 2,171,853 | 0.76% (0.50% - 1.07% |
| 40-49 years | 86 | 27% | 63 | 0.39% | 16,264 | 2,157,398 | 0.75% (0.54% - 1.02% |
| 50-59 years | 208 | 27% | 152 | 0.68% | 22,478 | 2,440,051 | 0.92% (0.68% - 1.23% |
| 60-64 years | 216 | 47% | 114 | 1.95% | 5,885 | 639,365 | 0.92% (0.66% - 1.27% |
| 65-69 years | 295 | 47% | 156 | 1.95% | 8,037 | 564,123 | 1.42% (1.03% - 1.97% |
| 70-79 years | 1015 | 47% | 538 | 3.86% | 13,937 | 523,291 | 2.66% (1.81% - 4.04% |
| 80+ years | 1330 | 58% | 559 | 20.38% | 2,741 | 275,081 | 1.00% (0.46% - 2.34% |
| April | | | | | | | |
| 20-29 years | 15 | 15% | 13 | 0.12% | 10,974 | 2,233,707 | 0.49% (0.34% - 0.71% |
| 30-39 years | 23 | 15% | 20 | 0.35% | 5,538 | 2,171,853 | 0.26% (0.17% - 0.36% |
| 40-49 years | 31 | 24% | 24 | 0.39% | 6,104 | 2,157,398 | 0.28% (0.20% - 0.39% |
| 50-59 years | 72 | 24% | 55 | 0.68% | 8,101 | 2,440,051 | 0.33% (0.24% - 0.45% |
| 60-64 years | 71 | 51% | 35 | 1.95% | 1,788 | 639,365 | 0.28% (0.20% - 0.38% |
| 65-69 years | 138 | 51% | 68 | 1.95% | 3,476 | 564,123 | 0.62% (0.44% - 0.84% |

| | Hospitalizations | Vaccination coverage | Unvaccinated hospitalizations[1] | Calculated risk of hospitalization[1] | Backcalculated number of cases[1] | Unvaccinated population[1] | Risk of COVID-19 infection[1,2] |
|---|---|---|---|---|---|---|---|
| 70-79 years | 435 | 51% | 213 | 3.86% | 5,522 | 523,291 | 1.06% (0.72% - 1.60% |
| 80+ years | 611 | 62% | 232 | 20.38% | 1,139 | 275,081 | 0.41% (0.19% - 0.97% |
| May | | | | | | | |
| 20-29 years | 6 | 14% | 5 | 0.12% | 4,441 | 2,233,707 | 0.20% (0.14% - 0.29% |
| 30-39 years | 15 | 14% | 13 | 0.35% | 3,654 | 2,171,853 | 0.17% (0.11% - 0.24% |
| 40-49 years | 14 | 23% | 11 | 0.39% | 2,793 | 2,157,398 | 0.13% (0.09% - 0.18% |
| 50-59 years | 39 | 23% | 30 | 0.68% | 4,446 | 2,440,051 | 0.18% (0.13% - 0.24% |
| 60-64 years | 56 | 51% | 27 | 1.95% | 1,410 | 639,365 | 0.22% (0.16% - 0.30% |
| 65-69 years | 57 | 51% | 28 | 1.95% | 1,436 | 564,123 | 0.25% (0.18% - 0.35% |
| 70-79 years | 220 | 51% | 108 | 3.86% | 2,793 | 523,291 | 0.53% (0.36% - 0.81% |
| 80+ years | 265 | 59% | 109 | 20.38% | 533 | 275,081 | 0.19% (0.09% - 0.46% |
| June | | | | | | | |
| 20-29 years | 2 | 0% | 2 | 0.12% | 1,721 | 2,233,707 | 0.08% (0.05% - 0.10% |
| 30-39 years | 6 | 0% | 6 | 0.35% | 1,700 | 2,171,853 | 0.08% (0.05% - 0.10% |
| 40-49 years | 4 | 32% | 3 | 0.39% | 705 | 2,157,398 | 0.03% (0.03% - 0.05% |
| 50-59 years | 17 | 32% | 12 | 0.68% | 1,711 | 2,440,051 | 0.07% (0.06% - 0.11% |
| 60-64 years | 15 | 58% | 6 | 1.95% | 324 | 639,365 | 0.05% (0.04% - 0.08% |
| 65-69 years | 26 | 58% | 11 | 1.95% | 561 | 564,123 | 0.10% (0.08% - 0.16% |
| 70-79 years | 52 | 58% | 22 | 3.86% | 566 | 523,291 | 0.11% (0.08% - 0.18% |
| 80+ years | 74 | 65% | 26 | 20.38% | 127 | 275,081 | 0.05% (0.02% - 0.13% |
| July | | | | | | | |
| 20-29 years | 3 | 25% | 2 | 0.12% | 1,937 | 2,233,707 | 0.09% (0.05% - 0.10% |
| 30-39 years | 6 | 25% | 4 | 0.35% | 1,275 | 2,171,853 | 0.06% (0.02% - 0.05% |
| 40-49 years | 4 | 33% | 3 | 0.39% | 694 | 2,157,398 | 0.03% (0.00% - 0.00% |
| 50-59 years | 18 | 33% | 12 | 0.68% | 1,785 | 2,440,051 | 0.07% (0.03% - 0.05% |
| 60-64 years | 12 | 57% | 5 | 1.95% | 265 | 639,365 | 0.04% (0.02% - 0.03% |
| 65-69 years | 21 | 57% | 9 | 1.95% | 464 | 564,123 | 0.08% (0.04% - 0.07% |
| 70-79 years | 78 | 57% | 34 | 3.86% | 869 | 523,291 | 0.17% (0.07% - 0.15% |
| 80+ years | 90 | 80% | 18 | 20.38% | 88 | 275,081 | 0.03% (0.02% - 0.09% |
| August | | | | | | | |
| 20-29 years | 8 | 21% | 6 | 0.12% | 5,440 | 2,233,707 | 0.24% (NA - NA |
| 30-39 years | 8 | 21% | 6 | 0.35% | 1,790 | 2,171,853 | 0.08% (NA - NA |
| 40-49 years | 12 | 34% | 8 | 0.39% | 2,052 | 2,157,398 | 0.10% (NA - NA |
| 50-59 years | 23 | 34% | 15 | 0.68% | 2,247 | 2,440,051 | 0.09% (NA - NA |

| | Hospitalizations | Vaccination coverage | Unvaccinated hospitalizations[1] | Calculated risk of hospitalization[1] | Backcalculated number of cases[1] | Unvaccinated population[1] | Risk of COVID-19 infection[1,2] |
|---|---|---|---|---|---|---|---|
| 60-64 years | 16 | 50% | 8 | 1.95% | 411 | 639,365 | 0.06% (NA - NA |
| 65-69 years | 23 | 50% | 12 | 1.95% | 591 | 564,123 | 0.10% (NA - NA |
| 70-79 years | 98 | 50% | 49 | 3.86% | 1,269 | 523,291 | 0.24% (NA - NA |
| 80+ years | 131 | 66% | 45 | 20.38% | 219 | 275,081 | 0.08% (NA - NA |
| October | | | | | | | |
| 20-29 years | 64 | 1% | 63 | 0.12% | 54,533 | 2,233,707 | 2.44% (1.70% - 3.51% |
| 30-39 years | 102 | 1% | 101 | 0.35% | 28,606 | 2,171,853 | 1.32% (0.88% - 1.88% |
| 40-49 years | 112 | 1% | 111 | 0.39% | 28,725 | 2,157,398 | 1.33% (0.96% - 1.82% |
| 50-59 years | 293 | 1% | 290 | 0.68% | 42,942 | 2,440,051 | 1.76% (1.30% - 2.37% |
| 60-64 years | 251 | 8% | 231 | 1.95% | 11,870 | 639,365 | 1.86% (1.35% - 2.59% |
| 65-69 years | 402 | 8% | 370 | 1.95% | 19,011 | 564,123 | 3.37% (2.44% - 4.69% |
| 70-79 years | 1242 | 8% | 1,143 | 3.86% | 29,602 | 523,291 | 5.66% (3.88% - 8.67% |
| 80+ years | 1504 | 14% | 1,293 | 20.38% | 6,346 | 275,081 | 2.31% (1.08% - 5.50% |
| November | | | | | | | |
| 20-29 years | 28 | 3% | 27 | 0.12% | 23,376 | 2,233,707 | 1.05% (0.73% - 1.50% |
| 30-39 years | 52 | 3% | 50 | 0.35% | 14,289 | 2,171,853 | 0.66% (0.44% - 0.94% |
| 40-49 years | 60 | 7% | 56 | 0.39% | 14,456 | 2,157,398 | 0.67% (0.49% - 0.93% |
| 50-59 years | 131 | 7% | 122 | 0.68% | 18,036 | 2,440,051 | 0.74% (0.55% - 1.01% |
| 60-64 years | 115 | 32% | 78 | 1.95% | 4,020 | 639,365 | 0.63% (0.48% - 0.92% |
| 65-69 years | 185 | 32% | 126 | 1.95% | 6,466 | 564,123 | 1.15% (0.86% - 1.66% |
| 70-79 years | 525 | 32% | 357 | 3.86% | 9,249 | 523,291 | 1.77% (1.27% - 2.84% |
| 80+ years | 610 | 37% | 384 | 20.38% | 1,886 | 275,081 | 0.69% (0.32% - 1.64% |
| December | | | | | | | |
| 20-29 years | 42 | 8% | 39 | 0.12% | 33,257 | 2,233,707 | 1.49% (1.03% - 2.14% |
| 30-39 years | 82 | 8% | 75 | 0.35% | 21,371 | 2,171,853 | 0.98% (0.65% - 1.39% |
| 40-49 years | 92 | 14% | 79 | 0.39% | 20,497 | 2,157,398 | 0.95% (0.68% - 1.30% |
| 50-59 years | 220 | 14% | 189 | 0.68% | 28,009 | 2,440,051 | 1.15% (0.85% - 1.55% |
| 60-64 years | 208 | 42% | 121 | 1.95% | 6,201 | 639,365 | 0.97% (0.73% - 1.40% |
| 65-69 years | 263 | 42% | 153 | 1.95% | 7,841 | 564,123 | 1.39% (1.05% - 2.01% |
| 70-79 years | 828 | 42% | 480 | 3.86% | 12,441 | 523,291 | 2.38% (1.70% - 3.79% |
| 80+ years | 1083 | 47% | 574 | 20.38% | 2,816 | 275,081 | 1.02% (0.48% - 2.46% |

[1]The vaccinated population is excluded from this analysis

[2]Between brackets are the low and high values used in the scenarios, based on a bootstrapped credible interval (2.5th and 97.5th percentile)

##### *Calculation risk of Myocarditis after infection*

A recent comprehensive study by the European Medicines Agency (EMA) estimated age-stratified incidence rates for myocarditis and pericarditis over the study period 2017-2019[7]. The background data presents the myocarditis/pericarditis incidence of the unvaccinated in the Netherlands:

| | **Incidence myocarditis/pericarditis** | |
|---|---|---|
| **Age-group (years)** | **Female** | **Male** |
| 20-29 | 0.006% | 0.036% |
| 30-39 | 0.041% | 0.055% |
| 40-49 | 0.024% | 0.045% |
| 50-59 | 0.021% | 0.041% |
| 60-69 | 0.028% | 0.046% |
| 70-79 | 0.012% | 0.037% |
| 80+ | 0.050% | 0.046% |

These data was used with the HR of 5.16 from a meta-analysis to assess the risk of myocarditis in post- COVID-19 individuals[8]. The risk difference, or the excess risk of infection-induced myocarditis, was estimated as follows:

| | **Incidence risk of myocarditis/pericarditis** | | **Excess risk of infection-induced myocarditis** | |
|---|---|---|---|---|
| **Age-group (years)** | **Female** | **Male** | **Female** | **Male** |
| 20-29 | 0.031% | 0.185% | 0.025% | 0.149% |
| 30-39 | 0.211% | 0.285% | 0.170% | 0.229% |
| 40-49 | 0.126% | 0.233% | 0.102% | 0.188% |
| 50-59 | 0.106% | 0.210% | 0.086% | 0.169% |
| 60-69 | 0.144% | 0.239% | 0.116% | 0.192% |
| 70-79 | 0.060% | 0.192% | 0.049% | 0.155% |
| 80+ | 0.257% | 0.238% | 0.207% | 0.192% |

For scenario analysis, data from Boehmer et al. was used, a cohort study from the Centers for Diseases Control and Prevention (CDC) on patients with at least one hospital-based encounter during March 2020 to Januari 2021[9]. Data was presented per age-group. The risk of the patients with COVID-19 was compared with the risk of patients without COVID, to estimate the adjusted myocarditis risk difference, by age, of myocarditis in infected individuals. However, age-groups in the model did not align with the age-groups in Boehmer et al., therefore, an average of the two adjacent age-groups was used:

| Excess risk of infection-induced myocarditis | |
|---|---|
| 18-29 years | 0.080% |
| 30-39 years | 0.067% |
| 40-49 years | 0.093% |
| 50-59 years | 0.137% |
| 60-64 years | 0.137% |
| 65-69 years | 0.160% |
| 70-79 years | 0.181% |
| 80+ years | 0.208% |

#### *Risk of Myocarditis after COVID-19 vaccination*

The incidence rate of vaccine-induced myocarditis/pericarditis (per dose) in individuals age 18-39 years was estimated from a self-controlled case series analysis was undertaken on hospital admissions for myocarditis or pericarditis in England from February 2021 to February 2022[10]. The study focused on individuals eligible to receive the mRNA vaccines (BNT162b2 or mRNA-1273) for boosting. The attributable risk for an admission for myocarditis 0 to 6 days post vaccine was 2.5 (95% confidence interval [CI], 2.0-2.8) and 4.5 (95%CI, 3.9-4.8) per million vaccinations of the BNT162b2 booster and mRNA-1273 booster vaccine, respectively[10].

#### *Length of stay, costs and QALY losses of myocarditis/pericarditis*

For the days of time loss of myocarditis/pericarditis after COVID-19 infection and vaccination, the length of stay from Dutch Statistics is used[11]. These days are used to estimate both the costs and QALYs lost. Costs per hospital day were obtained from the Dutch Costing Manual (€476/day, in 2014) and a QALY loss of 0.7 was assumed (per day=0.7/365)[12,13]:

| | Days of time loss | Costs per event (€) | QALYs lost |
|---|---|---|---|
| 18-29 years | 4.90 | 2,849 | 0.0094 |
| 30-39 years | 5.30 | 3,081 | 0.0102 |
| 40-49 years | 5.30 | 3,082 | 0.0102 |
| 50-59 years | 7.80 | 4,536 | 0.0150 |
| 60-64 years | 10.30 | 5,989 | 0.0198 |
| 65-69 years | 10.30 | 5,989 | 0.0198 |
| 70-79 years | 10.40 | 6,048 | 0.0199 |
| 80+ years | 10.40 | 6,048 | 0.0199 |

#### *Input for calculation indirect effects*

| Productivity loss | |
|---|---|
| Percentage in labor force[14] | % |
| Age group | |
| 0-17 years | |

| | |
|---|---|
| 18-29 years | 83.6% |
| 30-39 years | 87.8% |
| 40-49 years | 87.1% |
| 50-59 years | 83.0% |
| 60-64 years | 64.6% |
| 65-69 years | 23.0% |
| 70-79 years | 4.4% |
| 80+ years | 0.0% |

### Results

##### CE plane

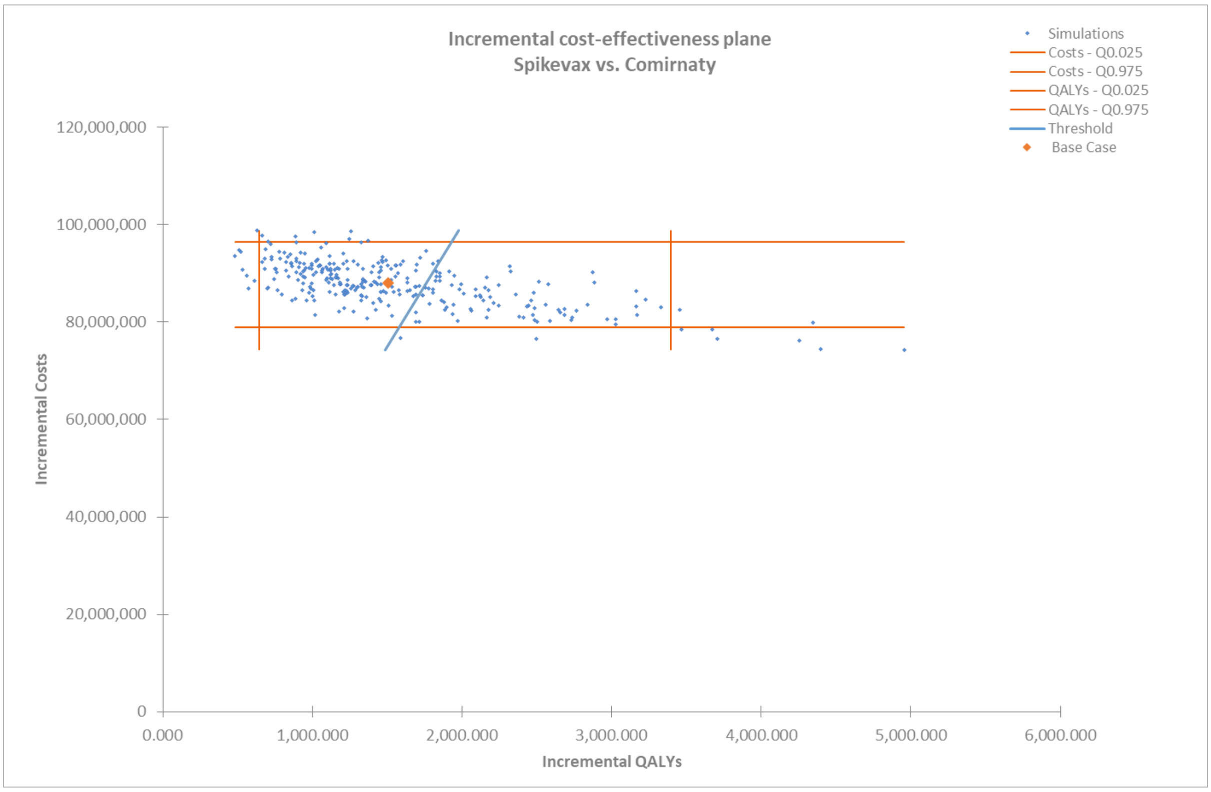
